# Internal pathway in Health Literacy among Chinese elementary school students: Serial Mediation Model Analysis

**DOI:** 10.1101/2025.08.13.25333503

**Authors:** Yuxing Wang, Jiayue Guo, Lizhu Liu, Mengyu Li, Ziqing Zan, Lili You

**Author notes:** Yuxing Wang, Jiayue Guo contributed equally.

## Abstract

The improvement of children’s health literacy is crucial for their physical and mental health development, and exploring the internal pathway of health literacy by age group is essential for improving the health literacy level at different age stages. We explored internal pathways within health literacy among elementary school students at different grade levels, utilizing a mediated effects model. A total of 3325 Chinese elementary students participated in a cross-sectional study between January to July 2023 using Health Literacy Questionnaire. IBM SPSS V.27 was used for descriptive statistics and correlation analysis of all variables. PROCESS SPSS was used to analyze chain-mediated effects. In grades 1-2, health motivation correlated with health participation through two paths: the mediating role of health skill, and the serial mediating roles of health knowledge and skill, presenting a partial mediating effect. In grades 3-4, health knowledge correlated with health participation through two paths: the mediating role of health motivation, and the mediating role of health skill, presenting a partial mediating effect. In grades 5-6, health knowledge correlated with health participation through one path: the mediating role of health motivation, presenting a total mediating effect. Our study suggests that elementary school students at different grade levels have different ways of improving health literacy because of the different pathways within health literacy. This provides new ideas for health education among children.

## Background

Health literacy (HL) has been defined as ‘the integration of skills, knowledge, and motivational drivers to access, understand, appraise, and apply health information for decision-making in healthcare, disease prevention, and health promotion contexts(Liu et al., 2020; Sørensen et al., 2012). For children, health literacy is similarly defined but within a developmental context that considers their capacity to cognitively understand health information, numeracy, and reading(Bröder et al., 2017a). Elementary school-aged children’s HL is critical because these younger children make everyday decisions about what to eat and how to play(Borzekowski, 2009). National and international surveys show that low HL is prevalent amongst children aged 10-24, ranging from 34% in the USA to 93.7% in China(Guo, 2018; Sansom-Daly et al., 2016). There is a clear social gradient in children’s HL (Fretian et al., 2020; Halfon et al., 2022; McDaid, 2016; Meherali et al., 2020; Pearce et al., 2019) and in the complex pathways from social determinants of health to health disparities, HL is not only a direct and independent determinant of health but also a mediating factor that influences the relationship between other social determinants of health (e.g., socioeconomic status) and health outcomes(Nutbeam & Lloyd, 2021).

With the increasing calls for more actions on addressing HL and inequalities (Bröder et al., 2017b), child HL interventions have emerged to bring about improvements in healthy behaviors(Fleary et al., 2018). HL is a key outcome of health education(Chang, 2008), the International Union for Health Promotion and Education (IUHPE) position statement on HL calls for global action to improve HL within populations through a systems approach(Bröder et al., 2018). Since its inception twenty years ago, the Health Promoting School (HPS) model (Langford et al., 2015; Young et al., 2013) has highlighted the importance of HL and there is evidence between the HPS approach with student health. Extensive research has shown that schools can provide inter-professional education and collaborative practice in promoting HL(Farokhi et al., 2018).

As a comprehensive complex of multi-dimensional abilities, HL requires the indispensable elements of knowledge, behavior, skills, and motivation, which complement each other(Van Boxtel et al., 2024). Despite this understanding of HL, there is limited consensus in health education research regarding on how to learn knowledge and master motivation for making appropriate health promotion decisions and actions of elementary school students(Bröder et al., 2017b). To be a “health literacy child” (Paakkari & Paakkari, 2012) describes HL as an individual competency which comprises theoretical and practical health knowledge, critical thinking, self-awareness, and citizenship, and corresponding learning conditions to develop students’ HL at school, so how knowledge, skill and motivation are viewed in relation to each other is also very important, however, previous researchers have preferred simple correlation analysis and ignored mediating mechanisms, the position and size of the roles these dimensions play in improving the children HL is not yet clear.

Previous studies have often viewed knowledge and skill (Knisel et al., 2020; Senahad et al., 2022; Truman et al., 2020) as a key component of children HL for health promotion and implementing health education(König et al., 2022; Ribeiro et al., 2023). So far, there are only a few studies that have examined the motivation within HL and there is a lack of empirical research on the mediating role of motivation. On the one hand, the existed children HL tools still have a lack of variety (Ormshaw et al., 2013) and often place too much emphasis on functional health literacy (knowledge or skill), and neglect the interactive and critical health literacy (motivation)(Fretian et al., 2020; Urstad et al., 2022). On the other, health education interventions often focus on quantifiable outcomes, such as knowledge acquisition, motivational enhancement effects are difficult to assess in the short term.

In this study, we argue that it is necessary to measure and improve HL within a theoretical framework to target effective interventions among different age groups. Currently, more than 20 theoretical frameworks have been proposed in the field of child HL(Bröder et al., 2017b). Here, we referred to Manganello’s HL framework (Manganello, 2008) as a guide, which views (1) self-efficacy and social support as upstream factors, (2) functional, interactive and critical as the construct of health literacy (Nutbeam, 2008), and (3) health behaviors, health service use as down-stream health outcomes. Besides, empirical studies have shown that personal self-efficacy (Guo et al., 2020) and health interests (Brown et al., 2007) are associated with child health literacy. Furthermore, we refer to Self-Determination Theory (SDT) and Information-Motivation-Behavioral Model (IMB). Thus, we used a Health Literacy Questionnaire which containing variables including self-efficacy, health knowledge, health behaviors, reading ability and numeracy as well as skills such as navigation and critical reasoning to explain the internal pathway of child HL.

In response to previous research findings, this study aims to identify the internal mediators of HL promotion. Specific research questions include: (1) what influences children HL; (2) whether knowledge or motivation acts as the mediator among HL promotion. We intend to conduct validation analyses with (i) motivation and skills as continuous mediating factors and (ii) knowledge and skills as continuous mediating factors, respectively; (3) should HL promotion strategies be tailored to different grade levels?

## Methods

### Study design and participants

This study was conducted from January to July 2023 using a multi-stage cluster sampling method to select the study subjects. At the first stage, in January and February 2023, four provinces spanning the southern, central, and northern regions of China were selected: Sichuan Province, Henan Province, Shanxi Province, and Beijing. Subsequently, one city was randomly selected from each of these provinces, namely Luzhou, Zhengzhou, Taiyuan. Then, within each selected city, one urban primary school and one rural primary school were chosen. In each of the selected schools, grades 1-2 were randomly sampled from each of the 1-2, 3-4, and 5-6 grade levels. The questionnaire survey was administered to all students within the selected sample classes and were completed by the students themselves independently. A total of 3325 valid questionnaires were collected. All participants provided informed consent.

### Health Literacy Questionnaire for Chinese elementary school students

Due to variations in cognitive levels and comprehension abilities among children of different age groups, this study developed three sub-questionnaires: HL-Grades 1-2 (Health Literacy Questionnaire for Grades 1-2), HL-Grades 3-4 (Health Literacy Questionnaire for Grades 3-4), and HL-Grades 5-6 (Health Literacy Questionnaire for Grades 5-6) (specific items of the three sub-questionnaires are provided in the supplementary file). The composition of the three questionnaires is the same, including (1) a general information questionnaire, including height and weight, self-assessment of health status, self-assessment of academic performance, self-assessment of personality, and ways of acquiring health-related knowledge, etc.; and (2) a measurement questionnaire, in which the internal dimensions are health knowledge (HK), health motivation (HM), health skill (HS) and health participation (HP).

The questionnaire has a total score of 100 points (scores for each internal dimension are presented in the supplementary file). All items have undergone validity and reliability testing using the Rasch model of Item Response Theory. The item reliability coefficient for all questionnaires is around 1, while the item separation indices are all exceeding 3, indicating good internal consistency for the questionnaires. The unidimensionality assumption holds true for all three questionnaires. For most items in the questionnaires, the average Infit MNSQ and Outfit MNSQ values fall between 0.5 and 1.5, suggesting a strong overall fit.

### Data analysis

The questionnaire item data were analyzed for their fit to the Rasch model using Winsteps (version 3.66.0; https://www.winsteps.com/index.htm).

IBM SPSS V.27 was used to complete the descriptive statistics and correlation analysis of all variables. If the data conformed to a normal distribution, the Pearson correlation coefficient was used to explore the relationship between health knowledge, health skill, health motivation, and health participation in primary school students’ health literacy.

PROCESS SPSS (version 4.1; developed by Andrew F. Hayes) was used to analyze chain-mediated effects. This analysis was conducted using the bias-corrected nonparametric percentile bootstrap method (model 6, with 5,000 replications and 95% confidence intervals) with a test level of α=0.05. This study investigated the effects of the mediator on the correlation between independent and dependent variables after controlling for demographic characteristics. Fully mediated occurs only when the effect of X (independent variable) on Y (dependent variable) is determined without controlling for the mediator. Partially mediated occurs when X exerts a significant effect on Y after controlling for the mediator.

#### Pathway 1: HK →HM →HS →HP

Model 1 examined the influence of health knowledge on health motivation. Model 2 explored the impact of health knowledge and motivation on health skill. Model 3 examined the combined effects of health knowledge, health motivation and health skill on health participation. Control variables such as age and gender were included as covariates in the models.

#### Pathway 2: HM →HK →HS →HP

Model 1 examined the influence of health motivation on health knowledge. Model 2 explored the impact of health motivation and health knowledge on health skill. Model 3 examined the combined effects of health motivation, health knowledge and health skill on health participation. Control variables such as age and gender were included as covariates in the models.

## Results

### Characteristics of participation

In this survey, there were 1070 students in grades 1-2, 1190 students in grades 3-4, and 1065 students in grades 5-6. Among different regions, Sichuan Province had the highest proportion, there was 74.02% in grades 1-2, 80.84% in grades 3-4, 60.66% in grades 5-6. The gender ratio of boys to girls was nearly equal, there was 47.38% female and 52.62% male in grades 1-2, 46.05% female and 53.95 male in grades 3-4, 48.26% female and 51.74% male in grades 5-6.

### Univariate analysis

Table 1 shows univariate analysis of HL composite score. For grades 1-2 and grades 3-4, there were significant differences in HL among region, daily outdoor activities, academic performance, character (*p*<0.05). For grades 5-6, there were significant differences in HL among region, gender, daily outdoor activities, easy tendency to catch a cold, academic performance, character (*p*<0.05).

**Table 1.**
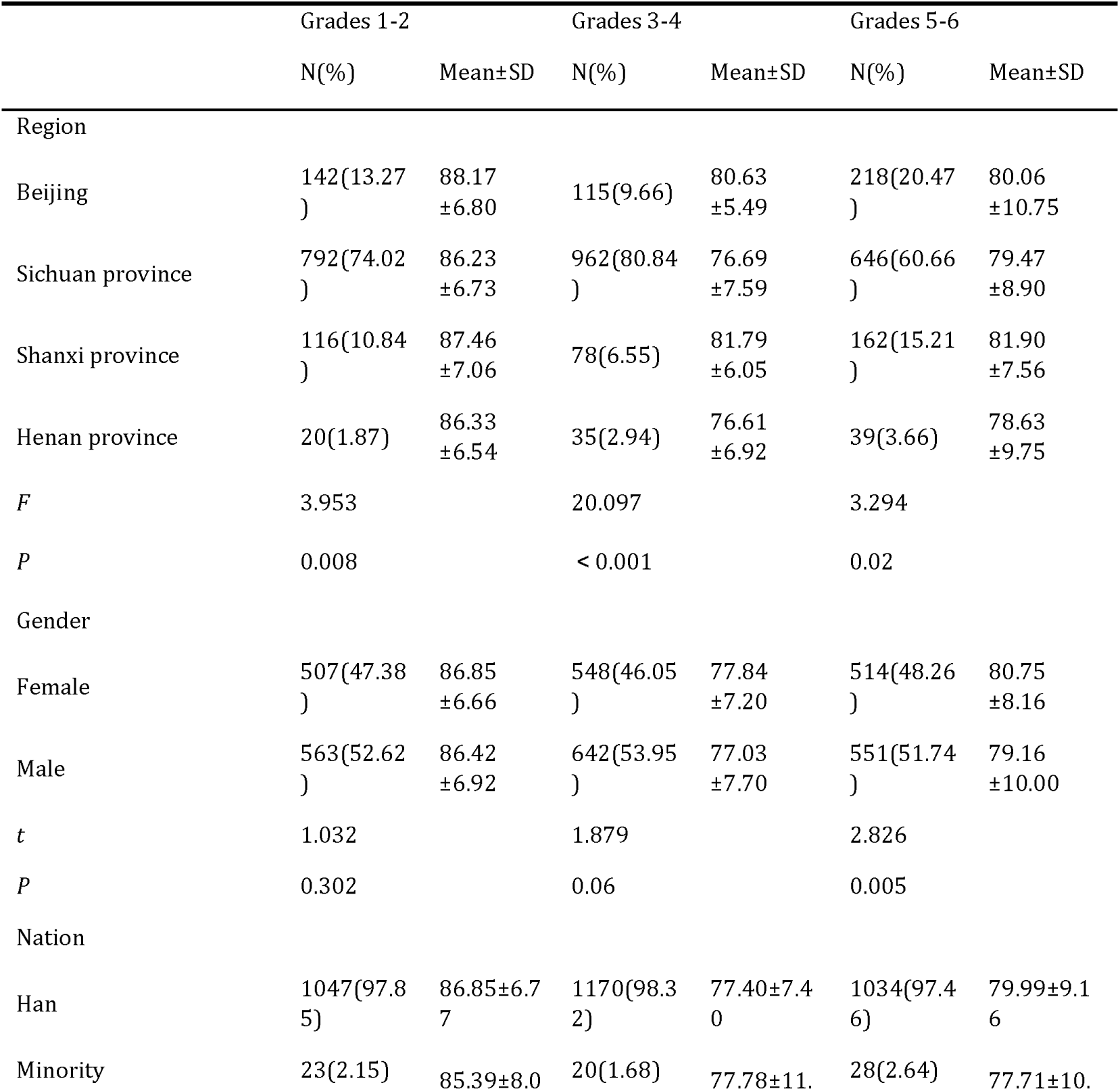
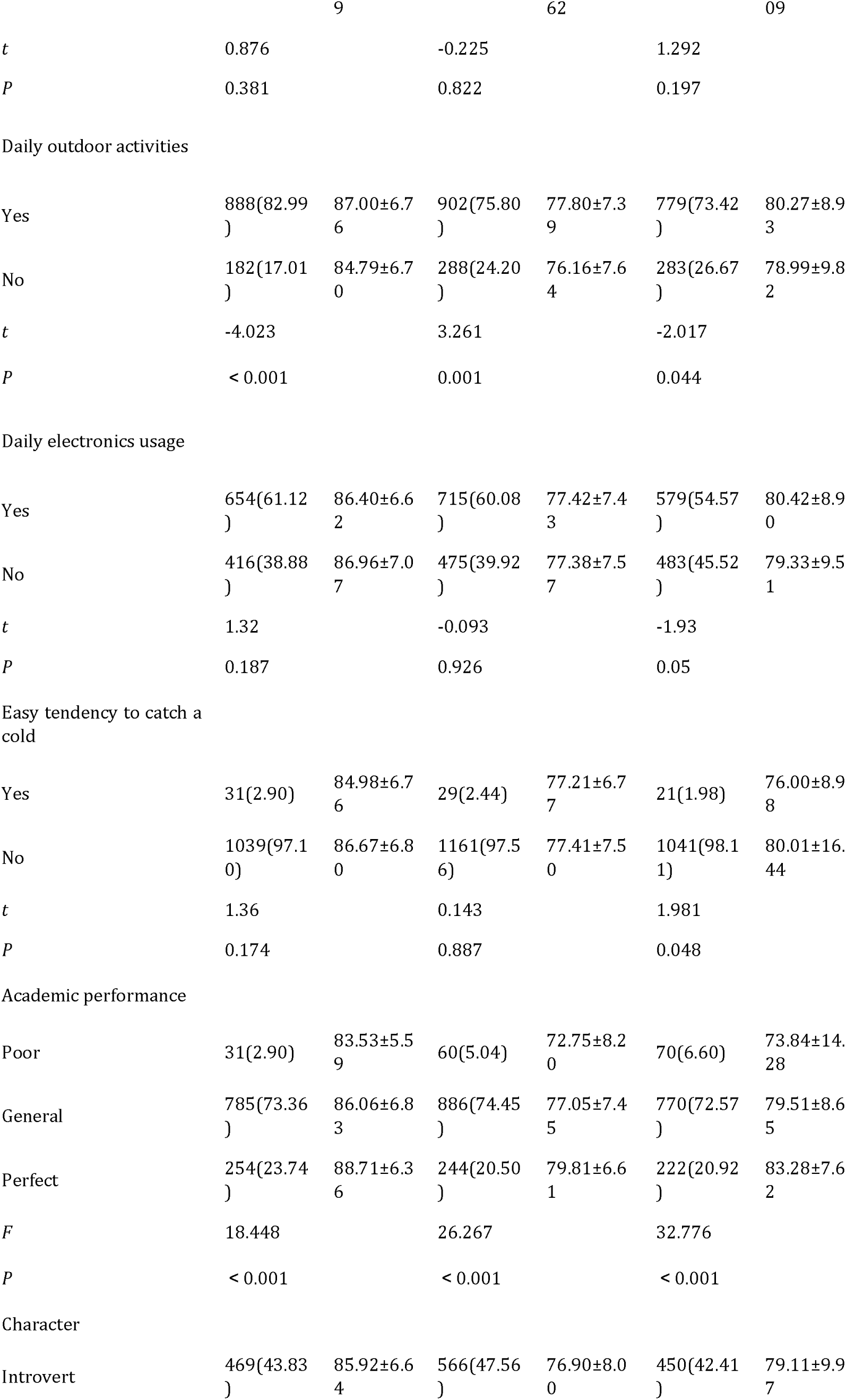
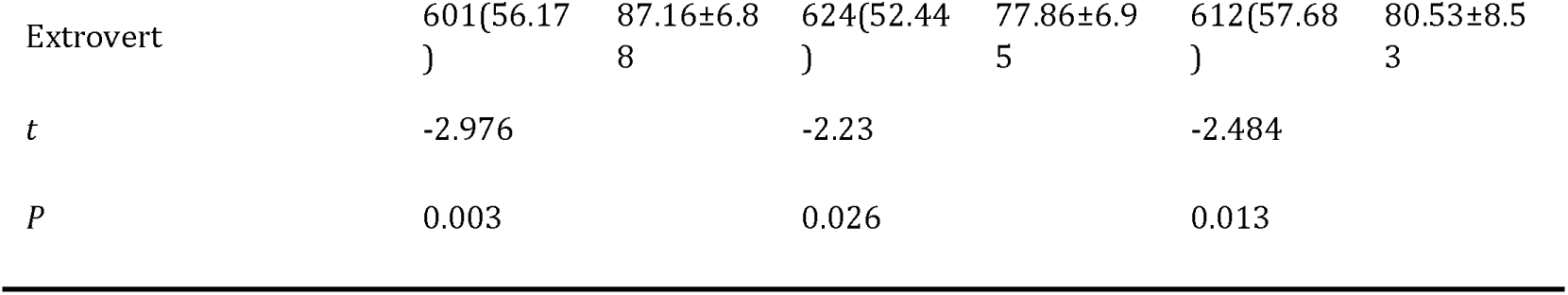
Univariate analysis of health literacy composite score.

|  | Grades 1-2 |  | Grades 3-4 |  | Grades 5-6 |  |
| --- | --- | --- | --- | --- | --- | --- |
|  | N(%) | Mean±SD | N(%) | Mean±SD | N(%) | Mean±SD |
| Region |  |  |  |  |  |  |
| Beijing | 142(13.27<br>) | 88.17<br>±6.80 | 115(9.66) | 80.63<br>±5.49 | 218(20.47<br>) | 80.06<br>±10.75 |
| Sichuan province | 792(74.02<br>) | 86.23<br>±6.73 | 962(80.84<br>) | 76.69<br>±7.59 | 646(60.66<br>) | 79.47<br>±8.90 |
| Shanxi province | 116(10.84<br>) | 87.46<br>±7.06 | 78(6.55) | 81.79<br>±6.05 | 162(15.21<br>) | 81.90<br>±7.56 |
| Henan province | 20(1.87) | 86.33<br>±6.54 | 35(2.94) | 76.61<br>±6.92 | 39(3.66) | 78.63<br>±9.75 |
| *F* | 3.953 |  | 20.097 |  | 3.294 |  |
| *P* | 0.008 |  | < 0.001 |  | 0.02 |  |
| Gender |  |  |  |  |  |  |
| Female | 507(47.38<br>) | 86.85<br>±6.66 | 548(46.05<br>) | 77.84<br>±7.20 | 514(48.26<br>) | 80.75<br>±8.16 |
| Male | 563(52.62<br>) | 86.42<br>±6.92 | 642(53.95<br>) | 77.03<br>±7.70 | 551(51.74<br>) | 79.16<br>±10.00 |
| *t* | 1.032 |  | 1.879 |  | 2.826 |  |
| *P* | 0.302 |  | 0.06 |  | 0.005 |  |
| Nation |  |  |  |  |  |  |
| Han | 1047(97.8<br>5) | 86.85±6.7<br>7 | 1170(98.3<br>2) | 77.40±7.4<br>0 | 1034(97.4<br>6) | 79.99±9.1<br>6 |
| Minority | 23(2.15) | 85.39±8.0 | 20(1.68) | 77.78±11. | 28(2.64) | 77.71±10. |

|  | 9 |  | 62 |  | 09 |  |
| --- | --- | --- | --- | --- | --- | --- |
| *t* | 0.876 |  | -0.225 |  | 1.292 |  |
| *P* | 0.381 |  | 0.822 |  | 0.197 |  |
| Daily outdoor activities |  |  |  |  |  |  |
| Yes | 888(82.99<br>) | 87.00±6.7<br>6 | 902(75.80<br>) | 77.80±7.3<br>9 | 779(73.42<br>) | 80.27±8.9<br>3 |
| No | 182(17.01<br>) | 84.79±6.7<br>0 | 288(24.20<br>) | 76.16±7.6<br>4 | 283(26.67<br>) | 78.99±9.8<br>2 |
| *t* | -4.023 |  | 3.261 |  | -2.017 |  |
| *P* | < 0.001 |  | 0.001 |  | 0.044 |  |
| Daily electronics usage |  |  |  |  |  |  |
| Yes | 654(61.12<br>) | 86.40±6.6<br>2 | 715(60.08<br>) | 77.42±7.4<br>3 | 579(54.57<br>) | 80.42±8.9<br>0 |
| No | 416(38.88<br>) | 86.96±7.0<br>7 | 475(39.92<br>) | 77.38±7.5<br>7 | 483(45.52<br>) | 79.33±9.5<br>1 |
| *t* | 1.32 |  | -0.093 |  | -1.93 |  |
| *P* | 0.187 |  | 0.926 |  | 0.05 |  |
| Easy tendency to catch a cold |  |  |  |  |  |  |
| Yes | 31(2.90) | 84.98±6.7<br>6 | 29(2.44) | 77.21±6.7<br>7 | 21(1.98) | 76.00±8.9<br>8 |
| No | 1039(97.1<br>0) | 86.67±6.8<br>0 | 1161(97.5<br>6) | 77.41±7.5<br>0 | 1041(98.1<br>1) | 80.01±16.<br>44 |
| *t* | 1.36 |  | 0.143 |  | 1.981 |  |
| *P* | 0.174 |  | 0.887 |  | 0.048 |  |
| Academic performance |  |  |  |  |  |  |
| Poor | 31(2.90) | 83.53±5.5<br>9 | 60(5.04) | 72.75±8.2<br>0 | 70(6.60) | 73.84±14.<br>28 |
| General | 785(73.36<br>) | 86.06±6.8<br>3 | 886(74.45<br>) | 77.05±7.4<br>5 | 770(72.57<br>) | 79.51±8.6<br>5 |
| Perfect | 254(23.74<br>) | 88.71±6.3<br>6 | 244(20.50<br>) | 79.81±6.6<br>1 | 222(20.92<br>) | 83.28±7.6<br>2 |
| *F* | 18.448 |  | 26.267 |  | 32.776 |  |
| *P* | < 0.001 |  | < 0.001 |  | < 0.001 |  |
| Character |  |  |  |  |  |  |
| Introvert | 469(43.83<br>) | 85.92±6.6<br>4 | 566(47.56<br>) | 76.90±8.0<br>0 | 450(42.41<br>) | 79.11±9.9<br>7 |
| Extrovert | 601(56.17<br>) | 87.16±6.8<br>8 | 624(52.44<br>) | 77.86±6.9<br>5 | 612(57.68<br>) | 80.53±8.5<br>3 |
| *t* | -2.976 |  | -2.23 |  | -2.484 |  |
| *P* | 0.003 |  | 0.026 |  | 0.013 |  |

### Mediation pathway Analysis

The bias-corrected percentile bootstrap analysis revealed different results among different grades. Region, gender, nation, daily outdoor activities, daily electronics usage, easy tendency to catch a cold, academic performance, character are the control variables.

#### Pathway1: HK →HM →HS →HP

In grades 1-2, the results showed that health knowledge did not significantly influence health participation. Thus, in this pathway, health motivation and health skill did not serve as the mediator.

In grades 3-4, the results presented a partial mediation effect, showing significant indirect effects of health motivation and health skill on the relationship between health knowledge on health participation. The total effect of health knowledge on health participation was 0.356 (SE = 0.047, 95% CI [0.264, 0.449]). The total direct effect and indirect effect of health knowledge on health participation was 0.311 (SE = 0.049, 95% CI [0.214, 0.407]), and 0.026 (SE = 017, 95% CI [0.008, 0.047]), respectively. The indirect effects followed two paths: HK→ HM→ HP (estimated effect = 0.006); HK →HS →HP (estimated effect = 0.020). The mediation effects of the two paths accounted for 19.23% and 30.77% of the total indirect effects, respectively. Figure 1 shows the standardized path coefficients of the proposed serial multiple mediation model, representing the indirect path coefficients between the variables.

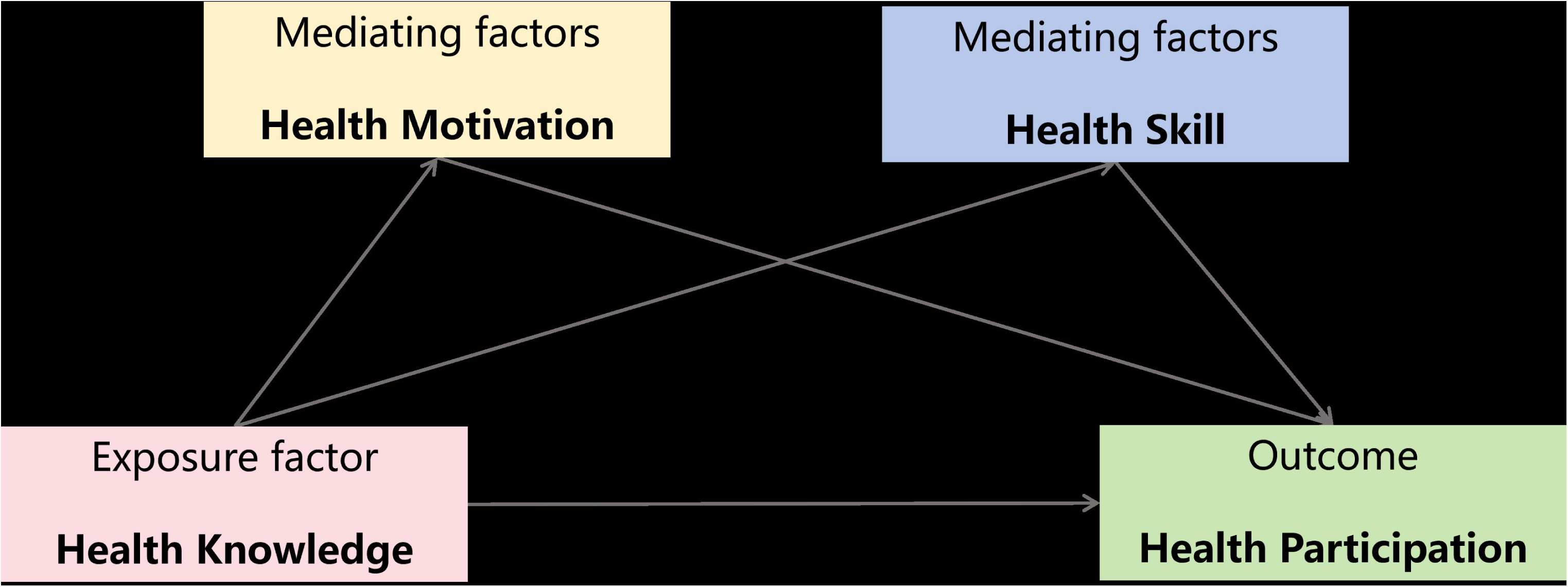

In grades 5-6, the results presented a complete mediation effect, showing significant indirect effects of health motivation and health skill on the relationship between health knowledge on health participation. The total effect of health knowledge on health participation was 0.068 (SE = 0.024, 95% CI [0.004, 0.022]). The total indirect effect of health knowledge on health participation was 0.047 (SE = 0.017, 95% CI [0.015, 0.083]). The indirect effects followed one path: HK→ HM→ HP (estimated effect = 0.028), which accounted for 121.28% of the total indirect effects. Figure 2 shows the standardized path coefficients of the proposed serial multiple mediation model, representing the indirect path coefficients between the variables.

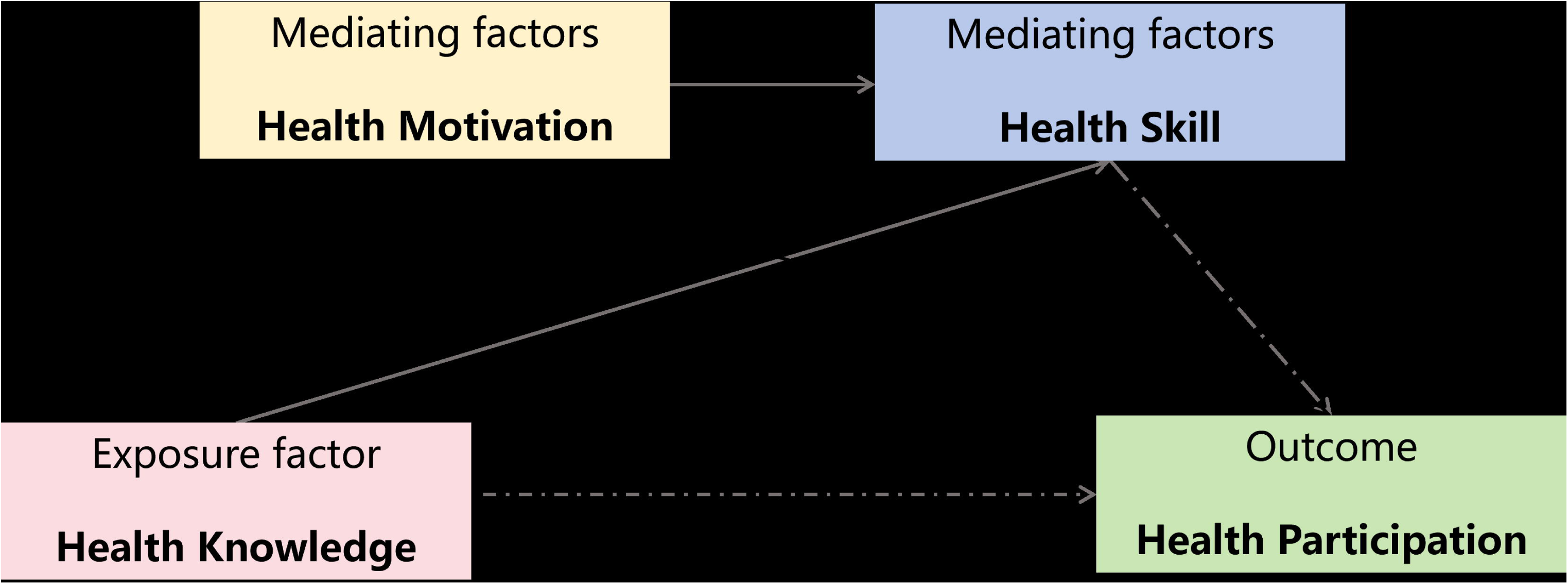

**Table 2.**
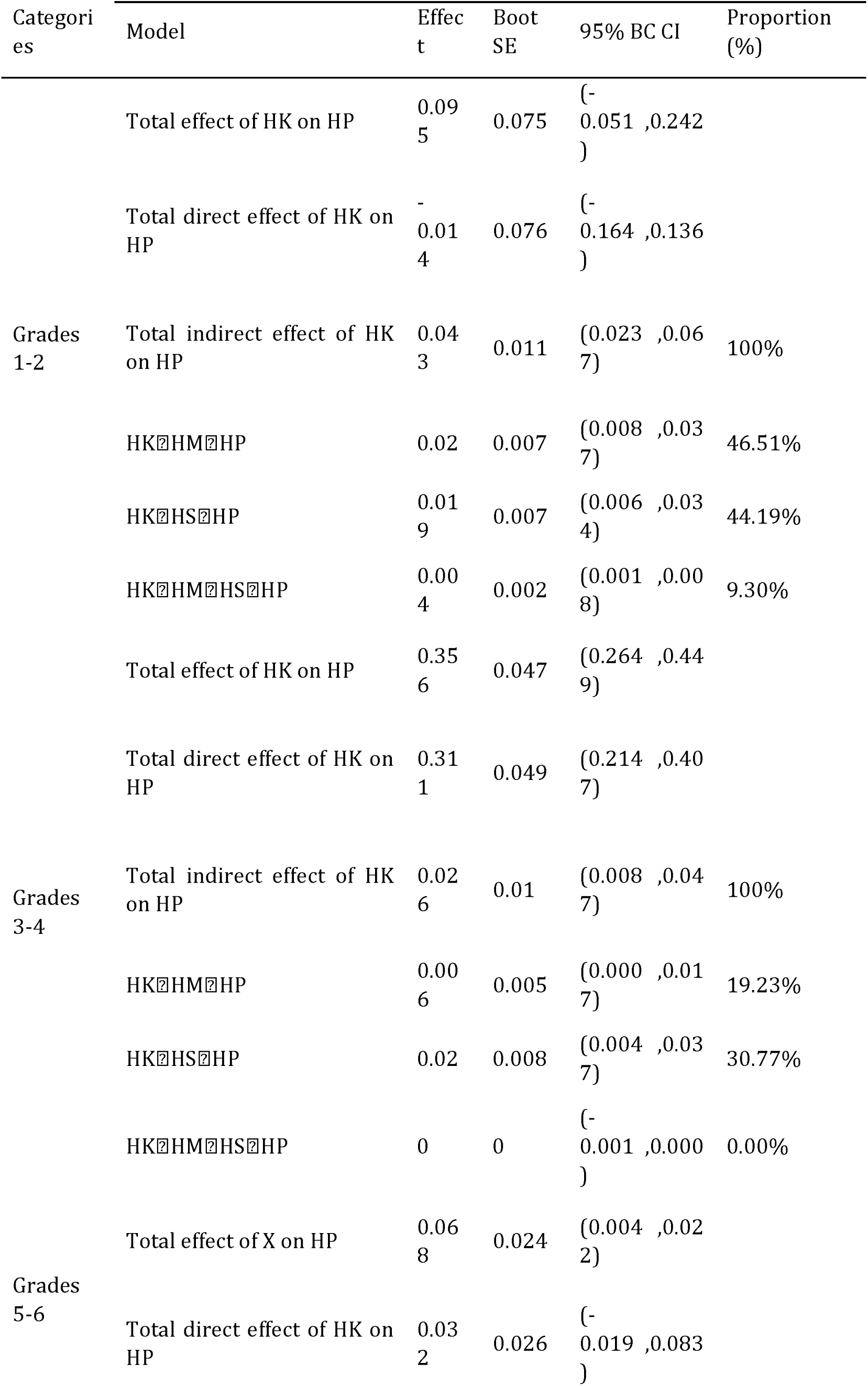
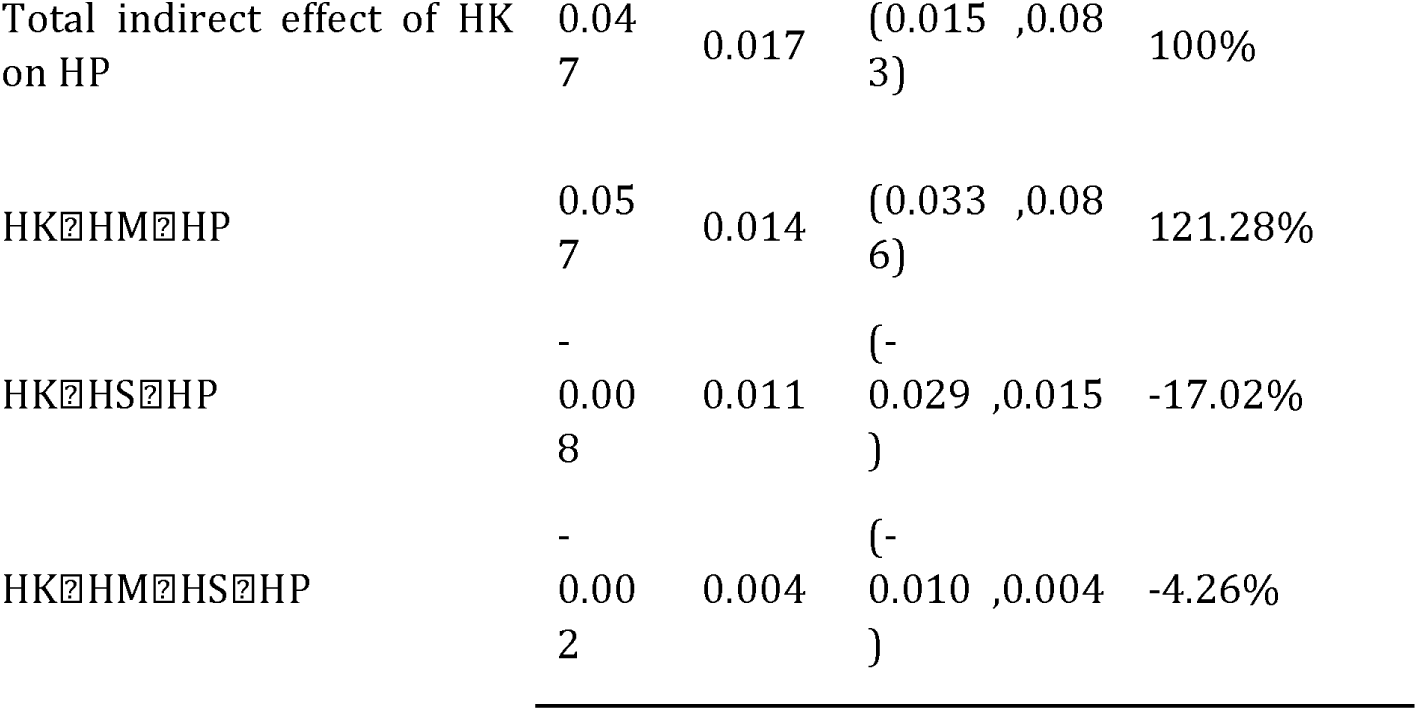
Indirect effects of health motivation and health skill. (pathway 1)

| Categori<br>es | Model | Effec<br>t | Boot<br>SE | 95% BC CI | Proportion<br>(%) |
| --- | --- | --- | --- | --- | --- |
| Grades<br>1-2 | Total effect of HK on HP | 0.09<br>5 | 0.075 | (-<br>0.051 ,0.242<br>) |  |
|  | Total direct effect of HK on<br>HP | -<br>0.01<br>4 | 0.076 | (-<br>0.164 ,0.136<br>) |  |
|  | Total indirect effect of HK<br>on HP | 0.04<br>3 | 0.011 | (0.023 ,0.06<br>7) | 100% |
|  | HK→HM→HP | 0.02 | 0.007 | (0.008 ,0.03<br>7) | 46.51% |
|  | HK→HS→HP | 0.01<br>9 | 0.007 | (0.006 ,0.03<br>4) | 44.19% |
|  | HK→HM→HS→HP | 0.00<br>4 | 0.002 | (0.001 ,0.00<br>8) | 9.30% |
| Grades<br>3-4 | Total effect of HK on HP | 0.35<br>6 | 0.047 | (0.264 ,0.44<br>9) |  |
|  | Total direct effect of HK on<br>HP | 0.31<br>1 | 0.049 | (0.214 ,0.40<br>7) |  |
|  | Total indirect effect of HK<br>on HP | 0.02<br>6 | 0.01 | (0.008 ,0.04<br>7) | 100% |
|  | HK→HM→HP | 0.00<br>6 | 0.005 | (0.000 ,0.01<br>7) | 19.23% |
|  | HK→HS→HP | 0.02 | 0.008 | (0.004 ,0.03<br>7) | 30.77% |
|  | HK→HM→HS→HP | 0 | 0 | (-<br>0.001 ,0.000<br>) | 0.00% |
| Grades<br>5-6 | Total effect of X on HP | 0.06<br>8 | 0.024 | (0.004 ,0.02<br>2) |  |
|  | Total direct effect of HK on<br>HP | 0.03<br>2 | 0.026 | (-<br>0.019 ,0.083<br>) |  |

|  |  |  |  |  |
| --- | --- | --- | --- | --- |
| Total indirect effect of HK on HP | 0.047 | 0.017 | (0.015 ,0.083) | 100% |
| HK→HM→HP | 0.057 | 0.014 | (0.033 ,0.086) | 121.28% |
| HK→HS→HP | -0.008 | 0.011 | (-0.029 ,0.015) | -17.02% |
| HK→HM→HS→HP | -0.002 | 0.004 | (-0.010 ,0.004) | -4.26% |

#### Pathway 2: HM →HK →HS →HP

In grades 1-2, the results presented a partial mediation effect, showing significant indirect effects of health knowledge and health skill on the relationship between motivation on health participation. The total effect of health motivation on health participation was 0.217 (SE = 0.046, 95% CI [0.126, 0.308]). The total direct effect and indirect effect of health knowledge on health participation were 0.180 (SE = 0.048, 95% CI [0.085, 0.275]), and 0.023 (SE = 0.010, 95% CI [0.005, 0.043]), respectively. The indirect effects followed two paths: HM →HS →HP (estimated effect = 0.021); and HM → HK →HS →HP (estimated effect = 0.003). The mediation effects of the two paths accounted for 91.30%, and 13.04% of the total indirect effects, respectively. Figure 3 shows the standardized path coefficients of the proposed serial multiple mediation model, representing the indirect path coefficients between the variables.

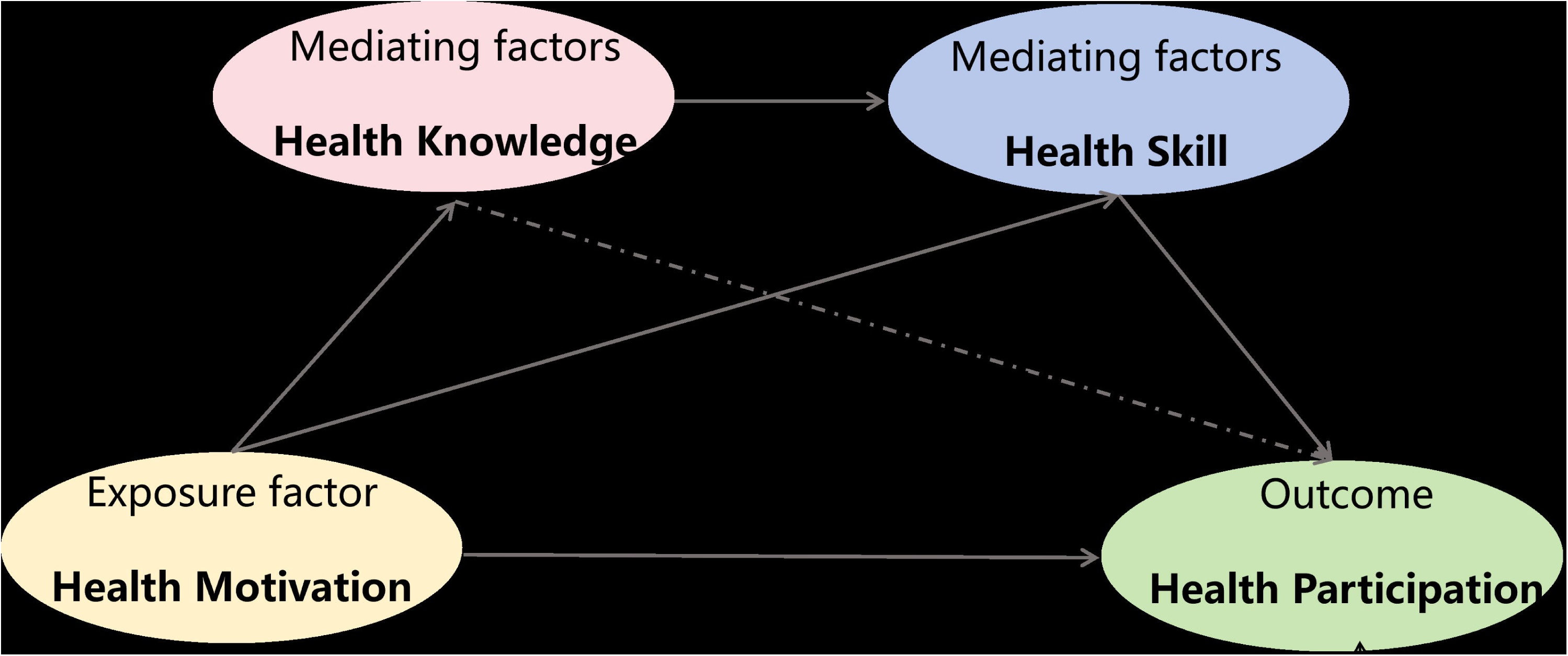

**Table 3.**
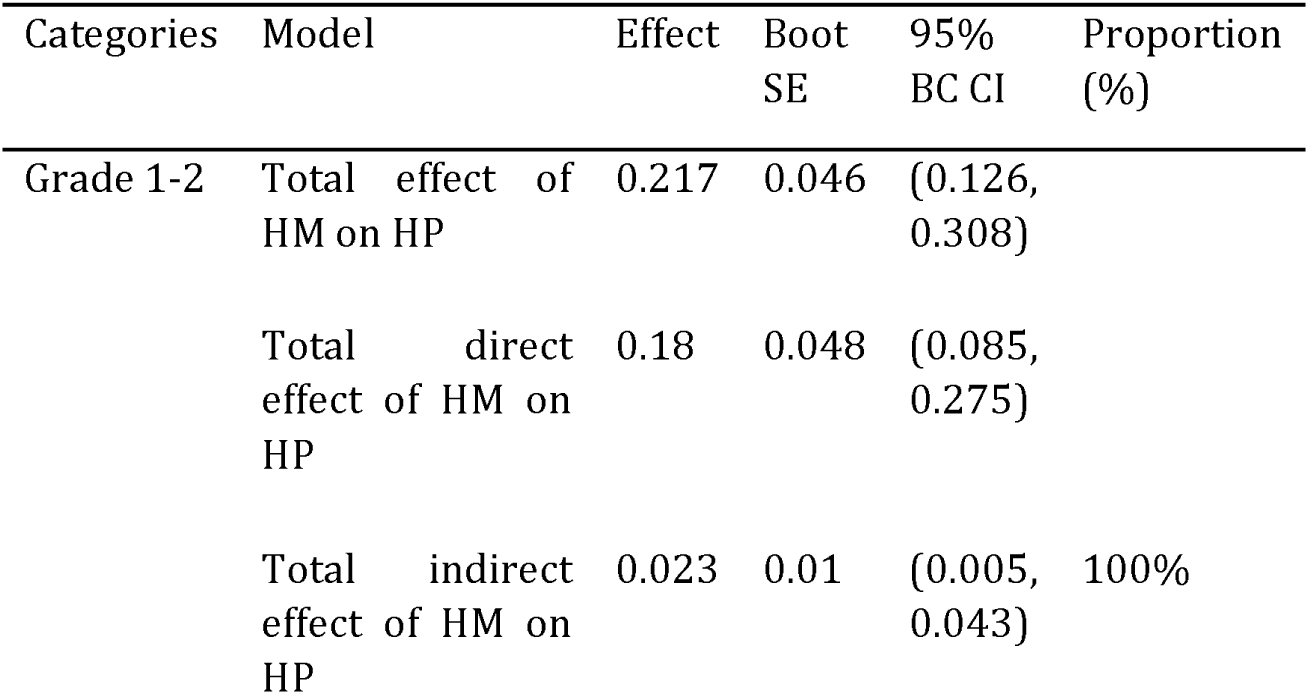
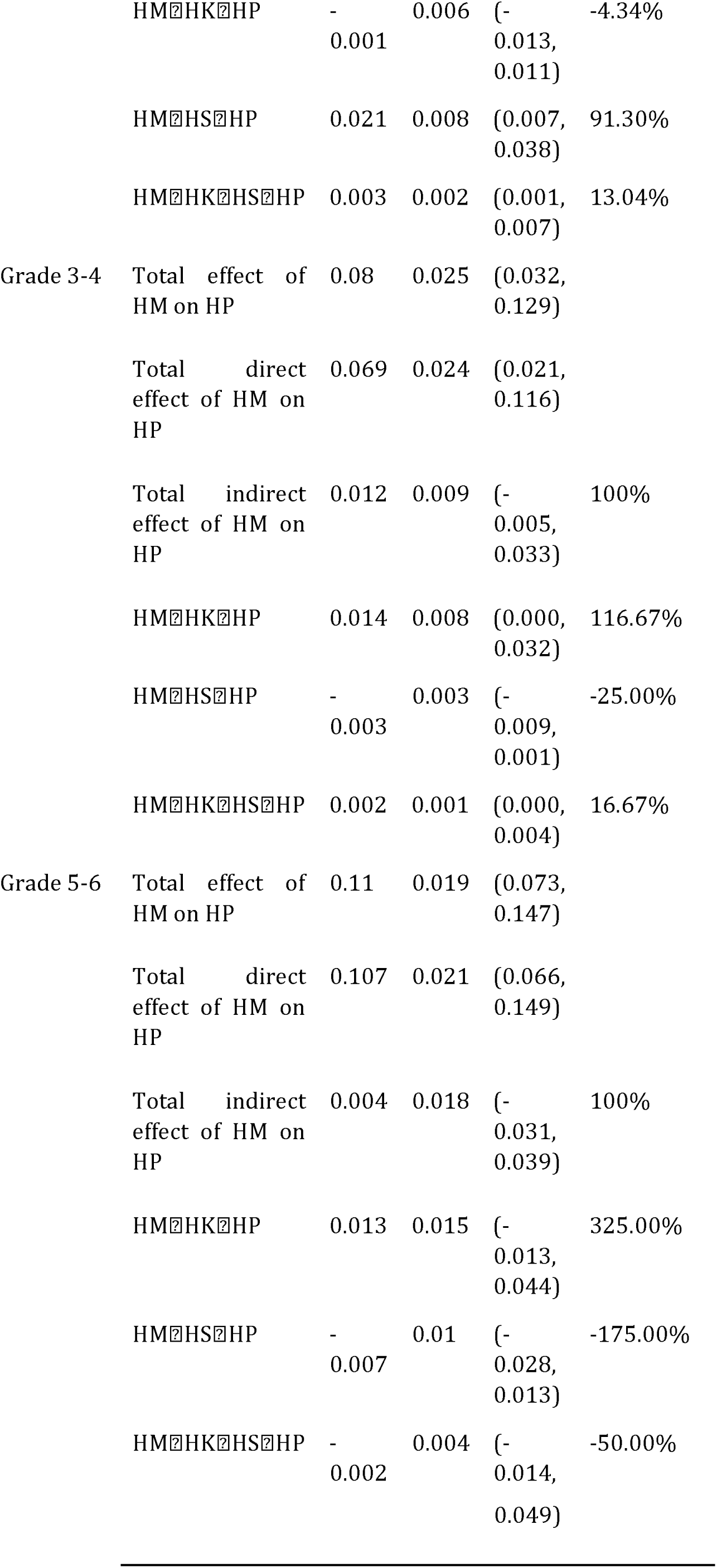
Indirect effects of health knowledge and health skill. (pathway 2)

In grades 3-4, the results showed that health motivation had a total and direct effect on participation but did not significantly have indirect effect on participation. Thus, in this pathway, health knowledge and health skill did not serve as the mediators. The total effect of health motivation on health participation was 0.080 (SE = 0.025, 95% CI [0.032, 0.129]). The total direct effect of health knowledge on health participation was 0.069 (SE = 0.024, 95% CI [0.021, 0.116]).

In grades 5-6, the results showed that health motivation had a total and direct effect on participation but did not significantly have indirect effect on participation. Thus, in this pathway, health knowledge and health skill did not serve as the mediators. The total effect of health motivation on health participation was 0.110 (SE = 0.019, 95% CI [0.073, 0.147]). The total direct effect of health knowledge on health participation was 0.107 (SE = 0.021, 95% CI [0.066, 0.149]).

## Discussion

We conducted an analysis of the factors influencing HL among students across different grade levels. Consistent with the literature, we found that there are differences in HL according to region(Senahad et al., 2022) and health condition(Adhikari et al., 2024). We found that gender demonstrated no statistically significant influence in grades 1-2 and grades 3-4, but as a statistically significant influence in grades 5-6. We also found that extrovert students with a higher score than introvert students and character had significant influence in HL. It may be attributed to extroverted students’ superior capacity in processing and retaining socially-interactive information, coupled with their heightened proactive participation in learning contexts.

Accumulating empirical evidence further indicates that inadequate health literacy correlates with adverse pediatric health outcomes, increased healthcare expenditures, and elevated mortality rates(Berkman et al., 2011; DeWalt & Hink, 2009).The IMB model posits that health knowledge serves as the foundational substrate, health motivation functions as the primary catalyst, and behavioral skills facilitate the actualization of health-promoting behaviors, thereby, we explored the pathway of HK-HN-HS-HP and found that the effect was not strong in grades 1-2. In conjunction with the SDT theory, we explored the pathway of HM-HK-HS-HP, in which health motivation provides the foundation, and learning health knowledge and improving health skills under the premise of health motivation further promote the enhancement of HL, and we found that the effect of this pathway was stronger for students in grades 1-2. This finding suggests that we should start from different perspectives to promote HL among elementary school students in different grade levels.

For grades 1-2, with health motivation as the independent variable, health knowledge and health skills as mediating variables, and health behavior as the dependent variable, this internal pathway has shown significant effects. This indicates that in grades 1-2, health knowledge and health skills play a crucial mediating role in health behavior. Students in grades 1-2 demonstrate an egocentric worldview perspective and are in the initial developmental phase of cognitive comprehension abilities(Borzekowski, 2009). Children within this developmental stage exhibit incomplete decentration and retain a predominantly egocentric orientation, wherein motivation serves as a pivotal mechanism of subjective agency, forming the foundational substrate for HL acquisition. The acquisition of health knowledge can further enhance health skills through practice, thereby facilitating the development of health participation.

For grades 3-4 and grades 5-6, the previous path did not show significant effects. Health knowledge acts as the independent variable, with health motivation and health skills serving as mediating variables, and health behavior as the dependent variable. This pathway has greater significance. The relationship between health knowledge and health behavior in grades 3-4 and grades 5-6 is mediated by health motivation and health skills. Furthermore, the study showed that health motivation played a significant role in HL internal pathways among every grade student. It may due to the unique health needs and characteristics: (1) Dependency on the environment: the younger the child, the greater their reliance on economic resources, social support, and parental health literacy(Borges et al., 2017; Miller et al., 2010; Ozturk Haney, 2020). (2) Development of stage specificity: children are in an important phase of growth, with significant variations in physical, emotional, and cognitive development across different ages and developmental stages(Yap et al., 2018). (3) Learning develops independence: Children aged 9–12 years old develop independence, while they transition from playful learning towards emphasizing academics in school and gaining information from peers and media(Van Boxtel et al., 2024). Students gradually transitioning from playful learning to school education, and gaining information from peers and various media(Van Boxtel et al., 2024; Velardo & Drummond, 2017). It is crucial to incorporate the unique health needs and developmental characteristics of children into the process of HL promotion(Van Boxtel et al., 2024). For different primary school student groups, specific and feasible HL strategies should be implemented to improve intervention efficiency.

Due to insufficient health education, students in underdeveloped countries exhibit lower productivity in terms of their health(Guszkowska & Dąbrowska-Zimakowska, 2022).Elementary school students’ HL was affected by health knowledge, self-motivation and school environment. it is important to broaden the perspectives and increase the coverage of targeted and purposeful health knowledge dissemination in the context of school. Other health literacy theoretical frameworks such as the social-ecological model(Wharf Higgins et al., 2009) and the HPS model(Langford et al., 2014) suggest that low HL is not only an individual’s issue, but results from interactions with the broader environment. Children are experiencing a life stage in which cognitive and social development processes take place(Bröder et al., 2020). Therefore, they are more likely to seek support from peers and parents when addressing health issues. In addition, children spend most of their daytime in schools where they learn health knowledge and health literacy skills(Nash et al., 2021). For grades 1-2 students, while fostering their healthy motivation and promoting their subjective initiative, their level of health knowledge and health skills should be improved. For students in grades 3-4 and 5-6 of elementary school, based on already possessing health knowledge, it is necessary to enhance their subjective initiative, so that they can actively acquire health-related skills, thereby further improving health behaviors, and ultimately raising the level of health literacy. To improve HL, a whole school approach is needed for the school-based intervention program that integrates strategies such as enhancing personal self-efficacy, improving social support, and creating supportive school environments(Paakkari & Paakkari, 2012).Therefore, we propose implementing differentiated motivation strategies across grade levels. For Grades 3-4 and 5-6, emphasis should be placed on peer education, incorporating group discussions and interactive formats in health knowledge learning. For Grades 1-2, the focus should be on instilling the concept of health responsibility, prioritizing it over knowledge acquisition. Elementary school student health education activities should be conducted to address the characteristics of different grades when researching HL, and optional HL interventions should be developed from multiple perspectives.

### Limitations

This study was a cross-sectional study and lacked follow-up on the longitudinal changes in the observed indicators. Future prospective studies or intervention studies related to health literacy and its internal pathway could be conducted to make the findings more robust.

## Conclusion

This study for the first time explored the internal pathway in health literacy (HL) among Chinese elementary school students and found that the four dimensions of HL (knowledge, motivation, skills, participation) are interdependent and collectively influence health behaviors. The analysis revealed that among younger students, health motivation directly affects health knowledge and skills, which in turn affect health participation; whereas among middle and high-grade students, health knowledge influences health participation through health motivation and skills. The study emphasizes the importance of implementing specific and feasible health literacy strategies tailored to different grade levels of elementary school students.

## Data Availability

All data produced in the present study are available upon reasonable request to the authors

## Abbreviations

(HL): Health Literacy;
(HPS): Health Promoting School;
(HK): Health knowledge;
(HM): Health Motivation;
(HS): Health Skill;
(HP): Health Participation;
(IMB): Information-Motivation-Behavioral Model;
(SDT): Self-Determination Theory

## Acknowledgements

This study was part of research funded by Science and Technology Innovation Project in Medicine and Health, Chinese Academy of Medical Sciences [grant number 2021-I2M-1-046]; National Natural Science Foundation of China [grant number 71904205]. We declare that these institutions played no role in the study design or in the analysis and interpretation of the data.

## Author Contributions

Yuxing Wang and Jiayue Guo reviewed the literature, performed the analyses, and wrote the draft of the manuscript. Lizhu Liu and Mengyu Li collected the data. The manuscript was reviewed and revised by Lizhu Liu and Lili You. All authors contributed to the interpretation of data and the final approved version.

## Supplementary File

Supplementary file is available at Health Promotion International online.

